# Live virus neutralizing antibodies against pre and post Omicron strains in food and retail workers in Québec, Canada

**DOI:** 10.1101/2023.09.03.23294976

**Authors:** Henintsoa Rabezanahary, Caroline Gilbert, Kim Santerre, Martina Scarrone, Megan Gilbert, Mathieu Thériault, Nicholas Brousseau, Jean-François Masson, Joelle N. Pelletier, Denis Boudreau, Sylvie Trottier, Mariana Baz

## Abstract

**Objectives:** To assess the neutralization activity pre and post Omicron BA.1 emergence in a unique cohort of 280 vaccinated restaurant/bar, grocery and hardware store workers in Québec, Canada.

**Methods:** Participants were recruited during the emergence of Omicron BA.1 variant. The neutralizing activity of participant sera was assessed by microneutralization assay.

**Results:** Serum neutralizing antibody (NtAb) titers of all participants against the ancestral SARS-CoV-2 strain was comparable with the response against Delta variant, however, their response was significantly reduced against Omicron BA.1, BA2, BA.2.12.1, BA.4 and BA.5. The neutralizing response of each group of workers was similar. Individuals who received 2 doses of vaccine had significantly reduced NtAb titers against all SARS-CoV-2 strains compared to those infected and then vaccinated (≥1 dose), vaccinated (≥2 doses) and then infected, or those who received 3 doses of vaccine. Participants vaccinated with 2 or 3 doses of vaccine and then infected had the highest NtAb titers against all SARS-CoV-2 strains tested.

**Conclusion:** We assessed for the first time the NtAb response in food and retail workers. Individuals infected after ≥2 doses of vaccine had the highest levels of NtAbs against Omicron BA.1, BA.2 and BA.5 variants and might be better protected against reinfection.

## Introduction

Since the beginning of the Coronavirus Disease 2019 (COVID-19) pandemic, the global response has faced new challenges including the emergence of new severe acute respiratory syndrome coronavirus 2 (SARS-CoV-2) variants of concern (VOC). In late 2020, the B.1.617.2 (Delta) variant was identified in India [1] and spread widely throughout the world. This variant harbors nine mutations in the viral spike (S) protein which alter biological characteristics such as binding affinity between the receptor binding domain (RBD) and the angiotensin-converting enzyme-2 (ACE2) receptor [2], as well as transmissibility and immune evasion [3]. In November 2021, Omicron (BA.1 sub-lineage of the B.1.1.529) variant emerged in South Africa [4] and rapidly spread around the world, becoming the dominant SARS-CoV-2 variant. Omicron BA.1 has acquired a large number of substitutions (>30), deletions and insertions in the spike protein and has been shown to escape protection conferred by vaccines and therapeutic monoclonal antibodies[5]. Since January 2022, several other Omicron sub-lineages have been detected such as BA.2, BA.2.12.1 (a variant of BA.2), BA.2.75, BQ.1.1, BA.4/5 and more recently XBB.1.5, XBB.1.16 and EG.5 [6]. Therefore, the global dynamic landscape of SARS-CoV-2 sub-genomes has become increasingly complex causing waves of infection in people with variable immunity induced by both infection and/or vaccination and have shown greatest evasion against parental or bivalent BA.1 or BA.4/5 mRNA-booster vaccines, explaining the rapid spreading of these new sub-lineages [7].

Neutralizing antibodies are crucial for virus clearance and are a major determinant of protection from infection in humans [8] and macaques challenged with SARS-CoV-2 [9]. In contrast with influenza infections, where a hemagglutination inhibition titer of 1:40 is thought to provide 50% protection from influenza infection [10], in the case of COVID-19, the role of NtAbs with regard to disease outcome remains undefined. It is also possible that neutralization is correlated with other immune responses such as T cell responses and B cell memory responses, which have also shown to contribute to protection [11]. Modeling studies have been used to estimate protective neutralization titer for COVID-19 [11], however, further studies and validations are needed.

Serological studies of SARS-CoV-2 antibodies have been conducted in several countries but have been focused on hospital and primary healthcare workers, blood donors, school children and staff and nursing homes [12–16]. Retail workers such as those working in grocery and hardware stores, restaurants and bars have been poorly or not studied.

In this study, we compared the neutralizing antibody levels in serum samples (n=280) against the ancestral SARS-CoV-2, Delta, Omicron BA.1, BA.2, BA.2.12.1, BA.4 and BA.5 strains, from a unique population composed by four groups of non-hospitalized (vaccinated, vaccinated and infected or vice versa) retail workers, including grocery and hardware stores, restaurants and bar workers in Quebec, Canada.

## METHODS

### Study participants

The 280 vaccinated participants were derived from a study which consisted of 304 food and retail workers who were recruited as part of a longitudinal study to assess the humoral and cellular responses to SARS-CoV-2 and its VOC in four groups of non-hospitalized retail workers [17]. Individuals were recruited in the Québec City area, specifically in the administrative regions of Capitale-Nationale and Chaudière-Appalaches, in Québec, Canada. The recruitment took place from October 2021 to May 2022, during the emergence of Omicron BA.1 variant, and individuals were classified into two groups: aged 18 to 59 and ≥60 years old. All participants provided information about vaccination and/or infection. The study was reviewed and approved by the CHU de Quebec-Université Laval Research Ethics Board (registration number 2021-5744). All experiments were performed in accordance with relevant guidelines and regulations. Adult volunteers were recruited at the *Centre Hospitalier Universitaire de Québec-Université Laval* (CHUL) in Quebec City. All participants provided informed written consent before enrolling. The study data is shared through the Maelstrom platform on a periodic basis as each visit is completed (https://www.maelstrom-research.org/study/cisacov).

### Sample Collection and Processing

Blood was collected in 6 mL tubes for serum, gently inverted, held at room temperature for 15-30 minutes and centrifuged at 1600 x *g* for 15 minutes. Aliquots of 1 mL of serum were transferred in cryovials and frozen at -20°C until used.

### Cells and viruses

Virus stocks used in this study were propagated in African green monkey kidney E6 cell line (Vero American Type Culture Collection, ATCC® CRL-1586^™^) or Calu-3 cells (ATCC-HTB-55) in 2% of Fetal Bovine Serum MEM and stored at -80°C until use. Live microneutralization assays were done in Vero E6 (ancestral SARS-CoV-2 and Delta variant) or Vero cells overexpressing transmembrane protease serine 2 (TMPRSS2, JCRB), (Omicron BA.1, BA.2, BA.2.12.1, BA.4 and BA.5 variants) cell lines. SARS-CoV-2/Québec City/21697/2020 strain (ancestral Wuhan-1 like SARS-CoV-2), was isolated from a clinical sample in March 2020 in Quebec City, Canada. Delta (SARS-CoV-2 VOC B.1.617.2) and Omicron sub-lineages BA.1 (SARS-CoV-2 VOC B1.1.529) and BA.2, BA.2.12.1, BA.4 and BA.5 were obtained from the National Microbiology Laboratory (NML), Public Health Agency of Canada.

### Live microneutralization assay

Microneutralization assays, the gold standard for evaluating virus NtAbs, were performed as previously described for influenza virus [18, 19] with some modifications. Briefly, sera from the participants were heat-inactivated (30 minutes at 56 °C), and serial two-fold dilutions were prepared from a 1:20 to 1:10,240 dilution of each sample. Equal volumes of serum and virus (100 TCID_50_ (50% tissue culture infectious dose) of each SARS-CoV-2 strain) were mixed and incubated for 60 min at room temperature. The residual infectivity of the virus-serum mixture was determined in Vero (ancestral and Delta variant) or Vero TMPRSS2, (Omicron BA.1 and BA.2, BA.2.12.1, BA.4 and BA.5 variants) cell lines using four wells for each dilution of serum. A virus back titration, positive controls (high, medium, and low titers) and negative controls were included in every experiment. Neutralizing antibody titer was defined as the reciprocal of the serum dilution that completely neutralized the infectivity of 100 TCID_50_ of each SARS-CoV-2 strain as determined by the absence of cytopathic effect on Vero or Vero TMPRSS2 cells at day 4 as previously described [18–20]. The neutralizing antibody titers are presented as geometric mean titre (GMT). These studies were performed in the Containment Level 3 (CL3) laboratory at the *CHU de Québec-Université Laval*.

### Statistical analysis

Sera with undetectable (<20) antibody titers were assigned an antibody titer of 10 for purposes of GMT calculations or statistical comparisons. Comparison between antibody titers against ancestral SARS-CoV-2, Delta and Omicron BA.1, BA2, BA.2.12.1, BA.4 and BA.5 variants were performed with Kruskal-Wallis one-way ANOVA followed by Dunn’s multiple comparison test, using GraphPad Prism 9.0 (GraphPad Software, Inc, San Diego, CA).

## RESULTS

We included 280 vaccinated participants; 105 (37-%), 133 (48%) and 42 (15%) worked in grocery stores, restaurants/bars or in hardware stores, respectively (Table 1). The median age of the 280 vaccinated participants was 41 years old (range 18-74) and 158 (56%) were females. Median body mass index was 26 kg/m^2^ (range 17-50). Chronic diseases (e.g., hypertension, diabetes, asthma, chronic lung, heart, kidney or liver disease, cancer, chronic blood disorder, immunosuppression, chronic neurological disorder), were present in 117 (42%) of the participants. Fifty-nine (21%) of the participants were smokers or e-cigarette users (vaping) (Table 1).

**Table 1:**
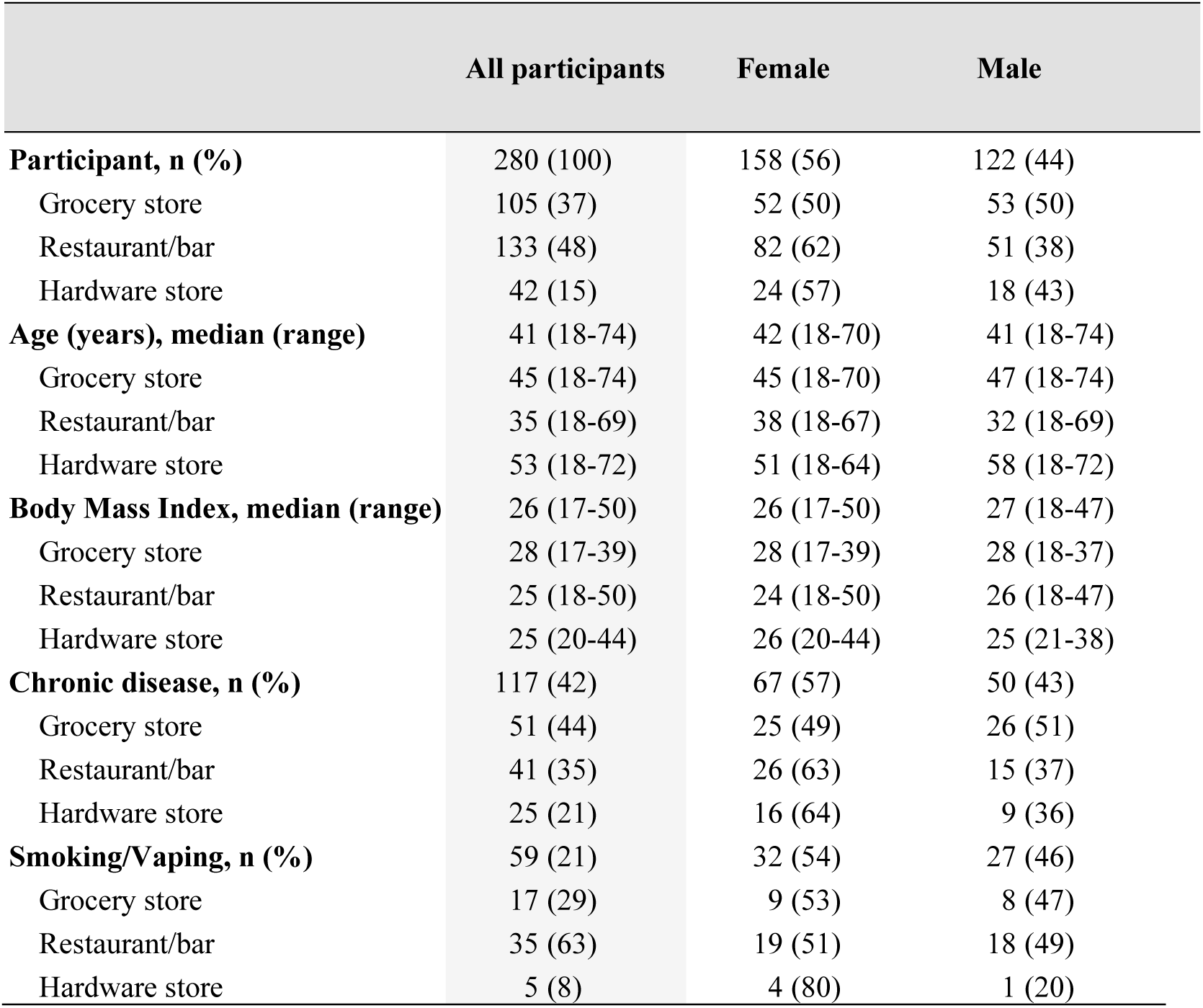
Demographic description and clinical characteristics of study participants.

|  | All participants | Female | Male |
| --- | --- | --- | --- |
| **Participant, n (%)** | 280 (100) | 158 (56) | 122 (44) |
| Grocery store | 105 (37) | 52 (50) | 53 (50) |
| Restaurant/bar | 133 (48) | 82 (62) | 51 (38) |
| Hardware store | 42 (15) | 24 (57) | 18 (43) |
| **Age (years), median (range)** | 41 (18-74) | 42 (18-70) | 41 (18-74) |
| Grocery store | 45 (18-74) | 45 (18-70) | 47 (18-74) |
| Restaurant/bar | 35 (18-69) | 38 (18-67) | 32 (18-69) |
| Hardware store | 53 (18-72) | 51 (18-64) | 58 (18-72) |
| **Body Mass Index, median (range)** | 26 (17-50) | 26 (17-50) | 27 (18-47) |
| Grocery store | 28 (17-39) | 28 (17-39) | 28 (18-37) |
| Restaurant/bar | 25 (18-50) | 24 (18-50) | 26 (18-47) |
| Hardware store | 25 (20-44) | 26 (20-44) | 25 (21-38) |
| **Chronic disease, n (%)** | 117 (42) | 67 (57) | 50 (43) |
| Grocery store | 51 (44) | 25 (49) | 26 (51) |
| Restaurant/bar | 41 (35) | 26 (63) | 15 (37) |
| Hardware store | 25 (21) | 16 (64) | 9 (36) |
| **Smoking/Vaping, n (%)** | 59 (21) | 32 (54) | 27 (46) |
| Grocery store | 17 (29) | 9 (53) | 8 (47) |
| Restaurant/bar | 35 (63) | 19 (51) | 18 (49) |
| Hardware store | 5 (8) | 4 (80) | 1 (20) |

Participants had been vaccinated with **≥** 1 dose of vaccine (messenger RNA (mRNA) vaccine BNT162b2 (Pfizer-BioNTech), mRNA-1273 (Moderna) or with viral vector vaccine ChAdOx1-S (AstraZeneca)), (Table 2). We analyzed participants regarding the number of vaccine doses received and if they were infected before or after vaccination and separated them in four groups: 1) infected and then vaccinated with 1-3 doses (n=16), 2) vaccinated with 2 doses (n=144), 3) vaccinated with 3 doses (n=84), and 4) vaccinated with 2-3 doses and then infected (n=36). We have considered participants in each group taking into consideration ≥7 days between last vaccination (or infection) and sample collection. The median delay between the last dose of vaccine or infection and the blood sample collection for group 1 was 315, 168 and 13 days for those who received one (n=2), two (n=11) or three doses of vaccine (n=3), respectively; for group 2 it was 157 days (n=144); for group 3 it was 48 days (n=84) and for group 4 it was 44 and 23 days for those who received two doses (n=28) and three doses of vaccine (n=8), respectively, (Table 2). The neutralizing antibody titers after the most recent vaccine dose or infection and the sample collection for each group stratified in intervals between 7-30 days, 31-60 days, 61-90 days and over 90 days is presented in Supplementary Figure 1.

**Table 2:**
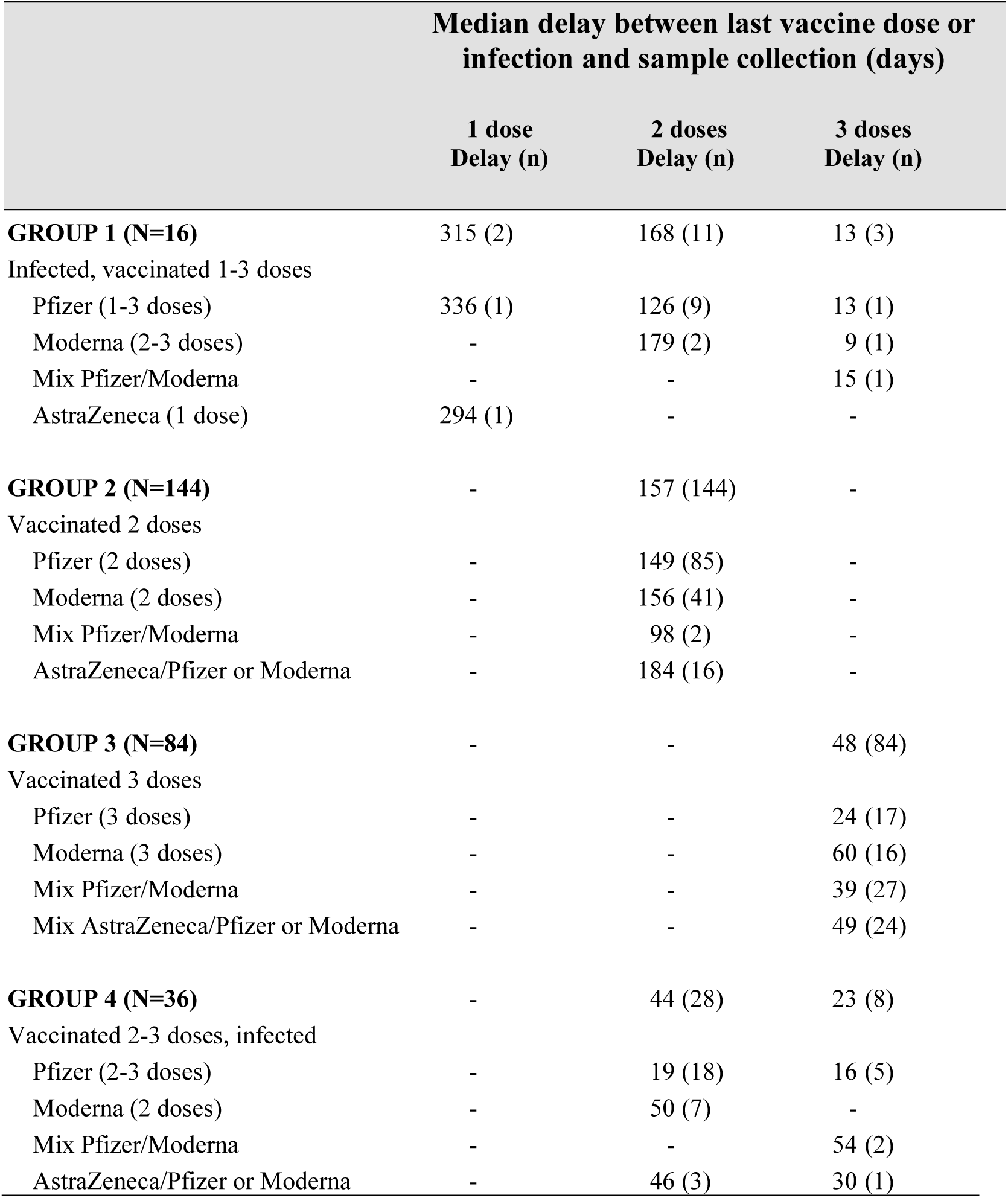
Median delay between last vaccine dose or infection and sample collection for groups 1 to 4.

| Median delay between last vaccine dose or infection and sample collection (days) |  |  |  |
| --- | --- | --- | --- |
|  | 1 dose<br>Delay (n) | 2 doses<br>Delay (n) | 3 doses<br>Delay (n) |
| **GROUP 1 (N=16)** | 315 (2) | 168 (11) | 13 (3) |
| Infected, vaccinated 1-3 doses |  |  |  |
| Pfizer (1-3 doses) | 336 (1) | 126 (9) | 13 (1) |
| Moderna (2-3 doses) | - | 179 (2) | 9 (1) |
| Mix Pfizer/Moderna | - | - | 15 (1) |
| AstraZeneca (1 dose) | 294 (1) | - | - |
| **GROUP 2 (N=144)** | - | 157 (144) | - |
| Vaccinated 2 doses |  |  |  |
| Pfizer (2 doses) | - | 149 (85) | - |
| Moderna (2 doses) | - | 156 (41) | - |
| Mix Pfizer/Moderna | - | 98 (2) | - |
| AstraZeneca/Pfizer or Moderna | - | 184 (16) | - |
| **GROUP 3 (N=84)** | - | - | 48 (84) |
| Vaccinated 3 doses |  |  |  |
| Pfizer (3 doses) | - | - | 24 (17) |
| Moderna (3 doses) | - | - | 60 (16) |
| Mix Pfizer/Moderna | - | - | 39 (27) |
| Mix AstraZeneca/Pfizer or Moderna | - | - | 49 (24) |
| **GROUP 4 (N=36)** | - | 44 (28) | 23 (8) |
| Vaccinated 2-3 doses, infected |  |  |  |
| Pfizer (2-3 doses) | - | 19 (18) | 16 (5) |
| Moderna (2 doses) | - | 50 (7) | - |
| Mix Pfizer/Moderna | - | - | 54 (2) |
| AstraZeneca/Pfizer or Moderna | - | 46 (3) | 30 (1) |

By the time of Omicron BA.1 emergence, 82% (86/105), 77% (103/133) and 81% (34/42) of grocery workers, restaurant/bar workers and hardware workers, respectively, were seropositive for ancestral SARS-CoV-2 and 80% (84/105), 77% (103/133) and 79% (33/42) were seropositive for the Delta variant. However, only 46% (48/105), 44% (59/103) and 38% (16/42) had cross-NtAbs against BA.1 variant.

Serum neutralizing antibody titers of all participants (n=280) against the ancestral SARS-CoV-2 strain was comparable with the response against Delta variant, with a geometric mean titer (GMT) of 76 (range 10-2032) and 71 (10-2560), respectively (Figure 1A). However, live NtAb response was reduced against the other variants tested, where the GMTs against BA.1, BA2, BA.2.12.1, BA.4 and BA.5 were 23 (range 10-1016), 26 (10-1016), 19 (10-320), 12 (10-80) and 17 (10-254), respectively. Thus, compared with the GMT of the ancestral SARS-CoV-2 strain, the neutralizing GMTs of the BA.1, BA2, BA.2.12.1, BA.4 and BA.5 were reduced by 3.3-, 3.0-, 4.0-, 6.3-, and 4.7-fold, respectively, (*p*<0.0001, Figure 1A). GMTs of BA.1 were 1.2-, 1.9-, and 1.4-fold higher compared to BA.2.12.1, BA.4 and BA.5, respectively, (*p*<0.0001, Figure 1A). No statistical difference was observed in the GMTs of each group of workers against each of the SARS-CoV-2 strain tested (Figure 1B). When we compared the GMTs against all SARS-CoV-2 strains of each of the groups (grocery and hardware stores and restaurant/bars), we observed a similar pattern than the whole cohort (Supplementary Figure 2).

**Figure 1.**
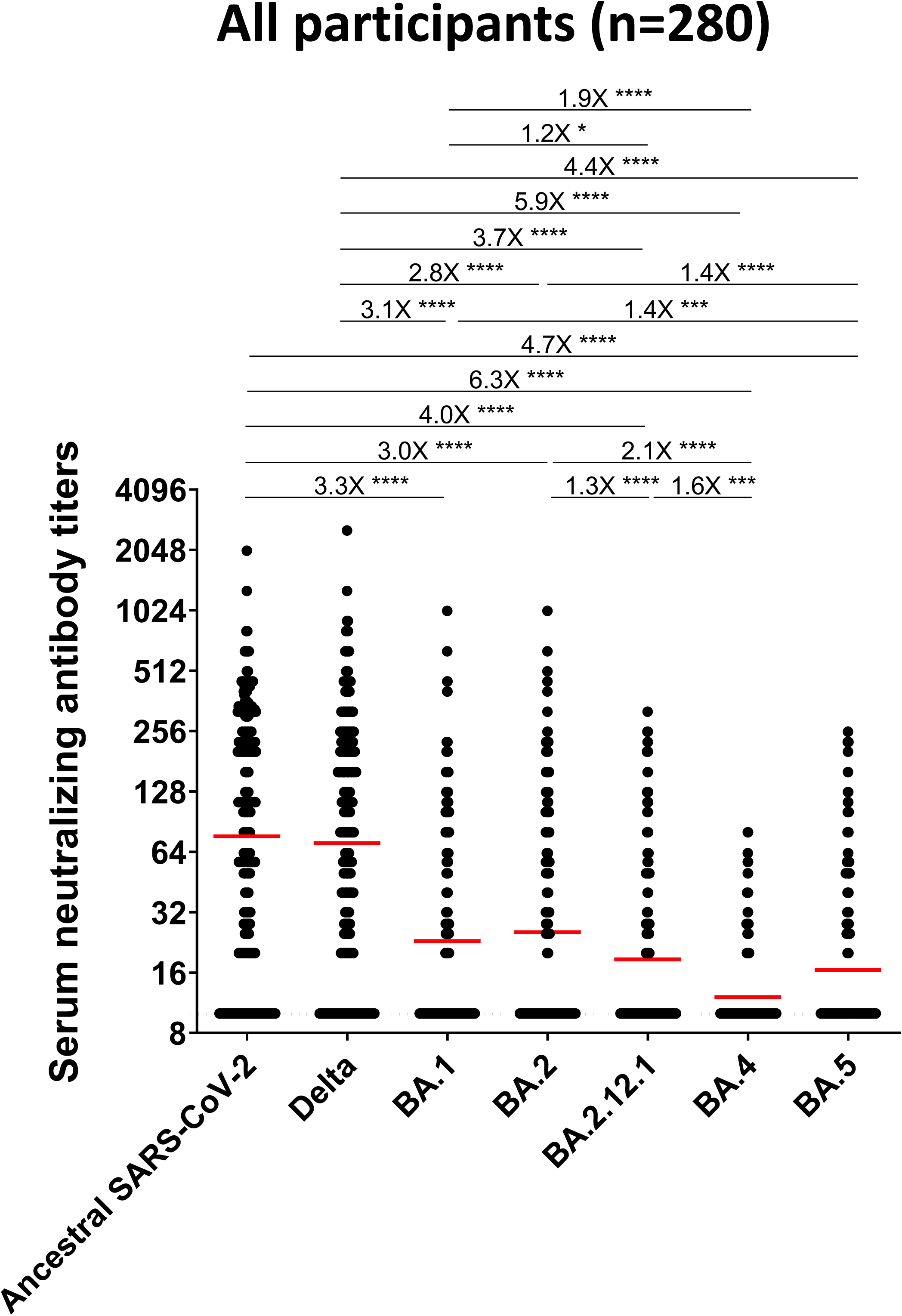
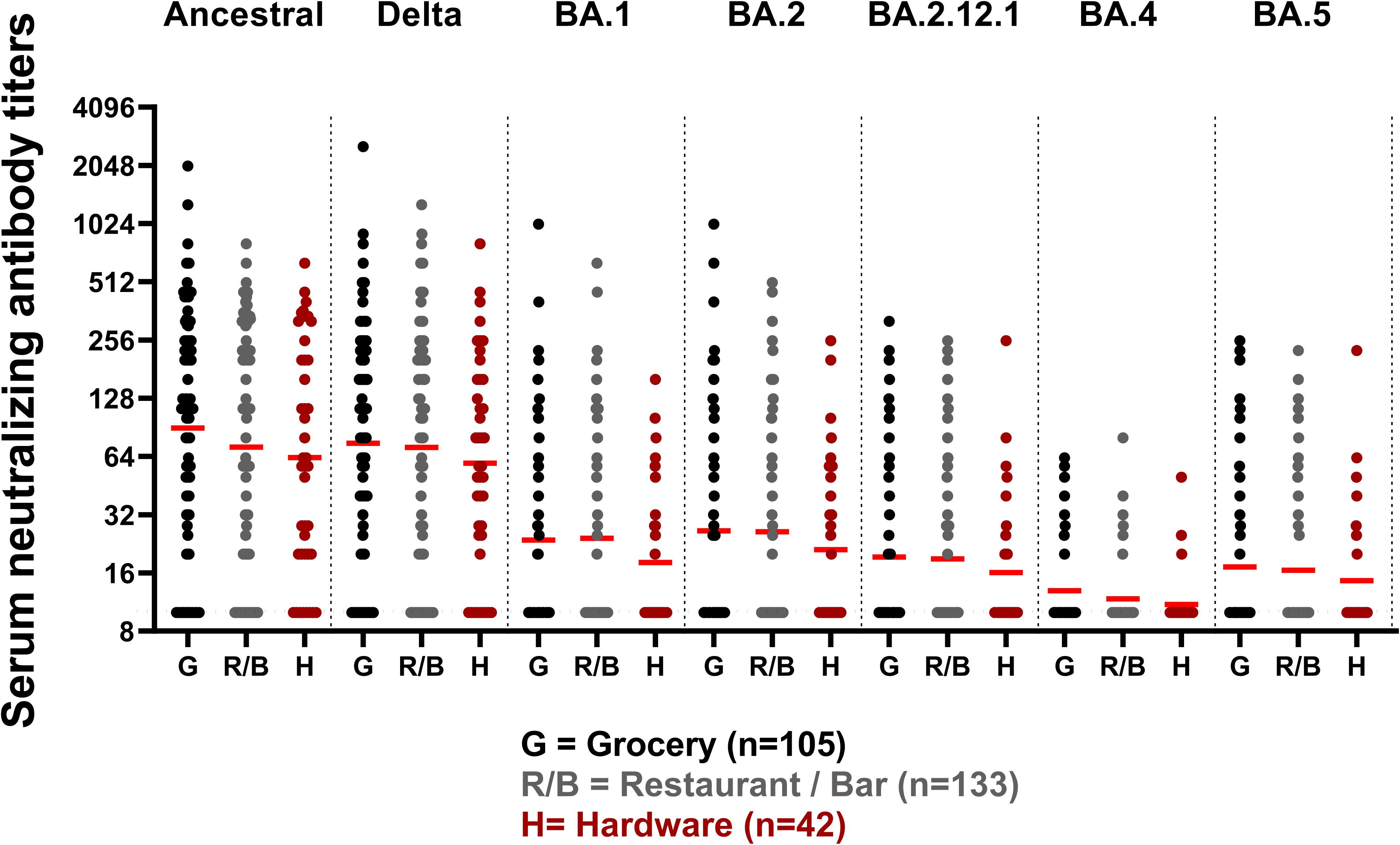
Comparison of live neutralizing antibody titers in participants after homologous or heterologous vaccination, infection followed by vaccination or vice-versa. Food and retail workers received two or three doses of mRNA vaccines (Pfizer, Moderna or AstraZeneca) or a combination of them. **(A)** Serum neutralizing antibody of all participants against ancestral SARS-CoV-2, Delta, BA.1, BA.2, BA.2.12.1, BA.4 and BA.5 strains. Each circle represents a single participant. Bars identify geometric mean titers of the group. The horizontal dashed line indicates the limit of detection for the neutralization assay (neutralizing titer of 10). The samples that did not neutralize SARS-CoV-2 at 1:20 serum dilution was given a neutralizing titer of 10 for graphic representation and statistical analysis. The fold-change of the geometric mean titer is denoted on the line. **(B)** Serum neutralizing antibody of participants from grocery stores (105), restaurants/bar (133) and hardware stores (42) against ancestral SARS-CoV-2, Delta, BA.1, BA.2, BA.2.12.1, BA.4 and BA.5 strains. Significance was assessed with Kruskal-Wallis one-way ANOVA followed by Dunn’s multiple comparison test (\**p*< 0.05; **** *p*< 0.0001).

We examined additional factors that may contribute to the NtAb response to vaccination or vaccination/infection (or vice-versa) in our food and retail workers cohort (all groups together) including sex and age, against all SARS-CoV-2 strains. We found a similar pattern in the humoral response in males (n=122) and females (n=158) between ancestral SARS-CoV-2 and the other variants. The GMTs against ancestral, Delta, BA.1, BA2, BA.2.12.1, BA.4 and BA.5 strains were 80, 69, 22, 24, 18, 12 and 16, respectively, for males (Figure 2A) and 74, 72, 23, 26, 19, 12, and 17, respectively, for females (Figure 2B). We did not find any significant difference between the age groups of <60 years (n=236, median 37 (range 18-59)) and ≥60 years (n=44, median 64 (range 60-73)) when we compared the GMT for the ancestral SARS-CoV-2 and Delta variants (GMT of 76 and 72 (10-1280) for both strains (p=0.99)). A 3.3-, 2.9-, 4.0-, 6.3- and 4.7-fold reduction was observed for BA.1, BA2, BA.2.12.1, BA.4 and BA.5 strains, respectively, when compared to the ancestral SARS-CoV-2, (Figure 2C) among participants of <60 years of age and a 3.8-, 3.3-, 4.4-, 6.7- and 4.7-fold reduction was observed against the same strains among participants of ≥60 years old (Figure 2D). No statistically significant differences were found in the GMTs against any SARS-CoV-2 strain in relation to the presence of chronic diseases and to smoking or vaping (Supplementary Figure 3). Interestingly, we found significantly higher GMTs against all strains except Delta in females with BMI ≥30 (Supplementary Figure 3C) as well as in smokers or vaping males, although in the latter group was only against Omicron BA.2.12.1 strain.

**Figure 2.**
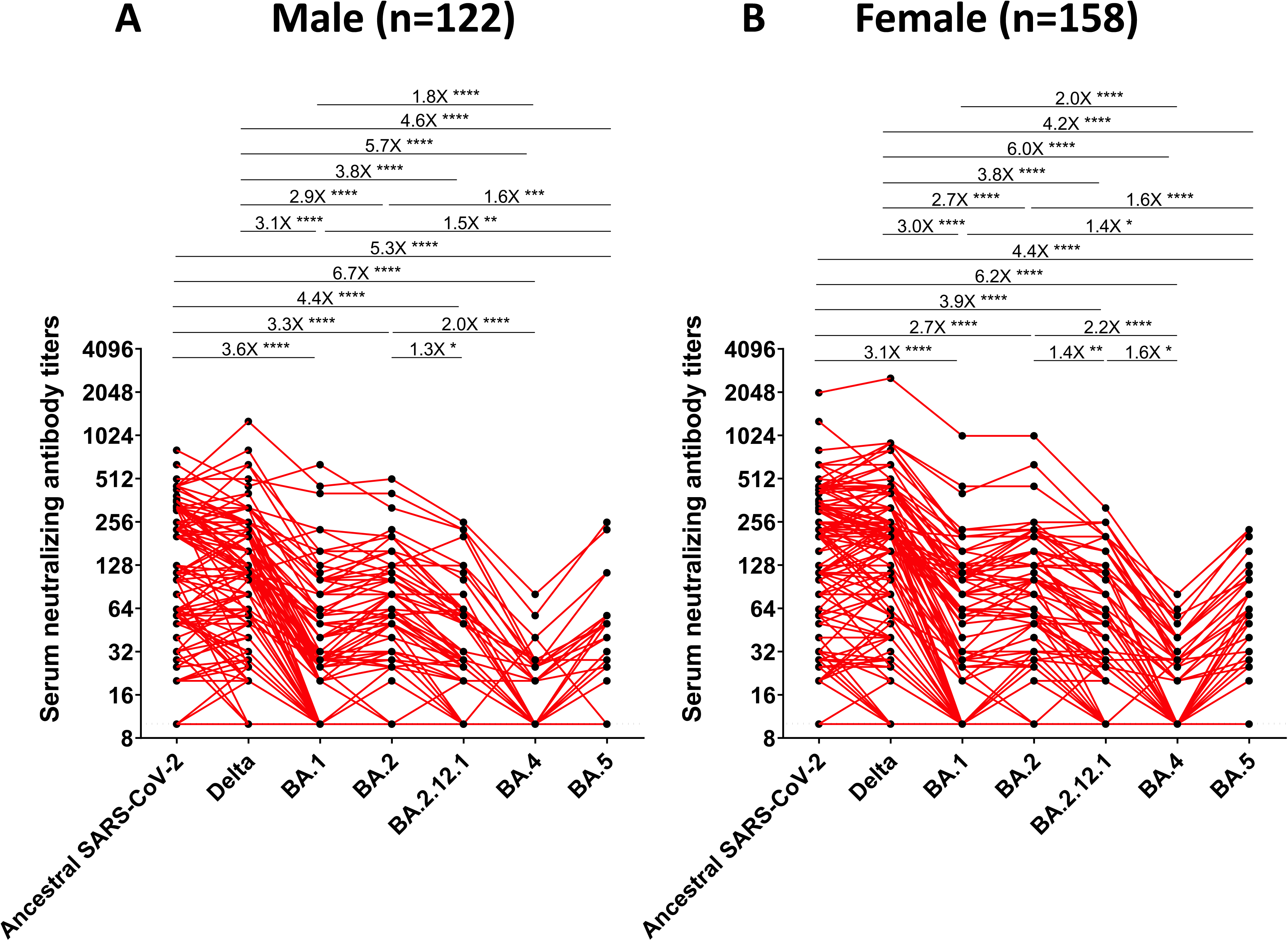
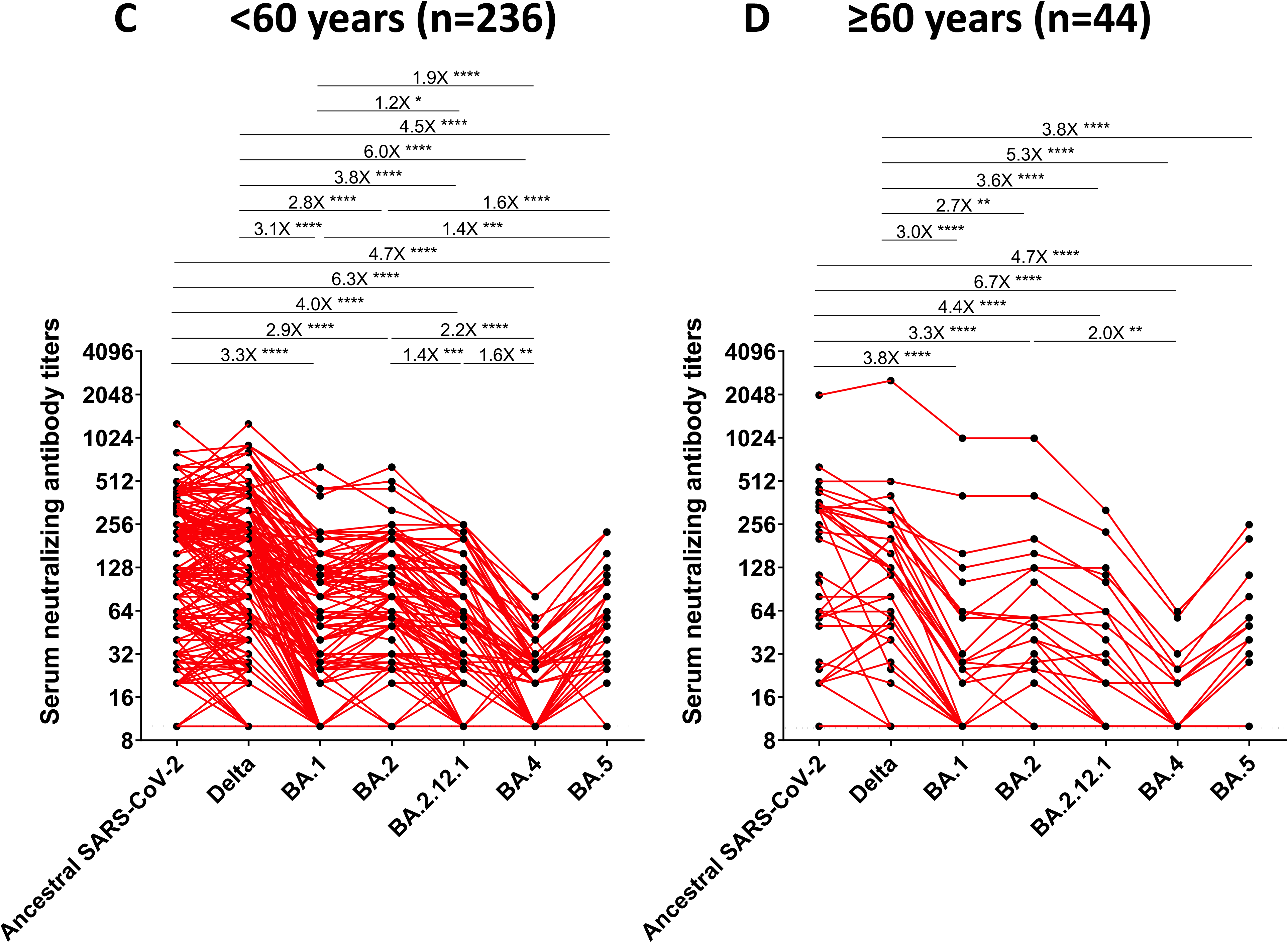
Neutralizing antibody titers in **(A)** males (n=122), **(B)** females (n=158), **(C)** <60 years (n=236, median 37 (range 18-59) and **(D)** ≥60 years (n=44, median 64 (60-73) against ancestral SARS-CoV-2, Delta, BA.1, BA.2, BA.2.12.1, BA.4 and BA.5 strains. Each circle represents a single participant. Bars identify geometric mean titers of the group. The horizontal dashed line indicates the limit of detection for the neutralization assay (neutralizing titer of 10). The samples that did not neutralize SARS-CoV-2 at 1:20 serum dilution was given a neutralizing titer of 10 for graphic representation and statistical analysis. The fold-change of the geometric mean titer is denoted on the line. Significance was assessed with Kruskal-Wallis one-way ANOVA followed by Dunn’s multiple comparison test (\**p* < 0.05; ** *p* < 0.01; *** *p* < 0.001; **** *p* < 0.0001).

The GMTs against the ancestral SARS-CoV-2 strain were statistically lower for group 2 (2 doses of vaccine, GMT=30) than for other groups: group 1 (infected, vaccinated with ≥ 1 dose, GMT=160, *p*=0.0021), group 3 (vaccinated with 3 doses, GMT=204, p<0.0001), group 4 (vaccinated with ≥ 2 dose and infected, GMT=221, p<0.0001) (Figure 3A). A similar profile was seen among the 4 groups against Delta variant. GMTs were lower in all groups against the other SARS-CoV-2 Omicron variants (BA.1, BA.2, BA.2.12.1, BA.4 and BA.5). Of note, the GMT of participants who were vaccinated with 2 or 3 doses of vaccine and then infected (group 4) had consistently higher NtAb titers against all variant tested, although a statistical significance was not always observed (Figure 3A). A similar trend was found for the other SARS-CoV-2 variants. Figure 3B shows the NtAb titers against each of the SARS-CoV-2 strains tested for participants of the 4 groups, each subgroup representing the type of vaccine regimen they have received. No significant differences were observed in the GMTs of the different subgroups against the ancestral SARS-CoV-2, except for those individuals who were vaccinated with 2 doses of Moderna (GMT=43) versus those who received AstraZeneca/Pfizer or AstraZeneca/Moderna (GMT=15), where a 2.9-fold reduction was observed (*p* = 0.01). Regarding the Delta variant, the combination of 2 doses of Pfizer (*p* = 0.03) or Moderna (*p* = 0.0026) or 3 doses of Pfizer (*p* = 0.0157) showed better response than a mixed schedule containing the AstraZeneca vaccine. Similar patterns were seen against BA.1 and BA.2 variants, but no significant differences were found when analyzing against BA.2.12.1, BA.4 and BA.5 (Figure 3B).

**Figure 3.**
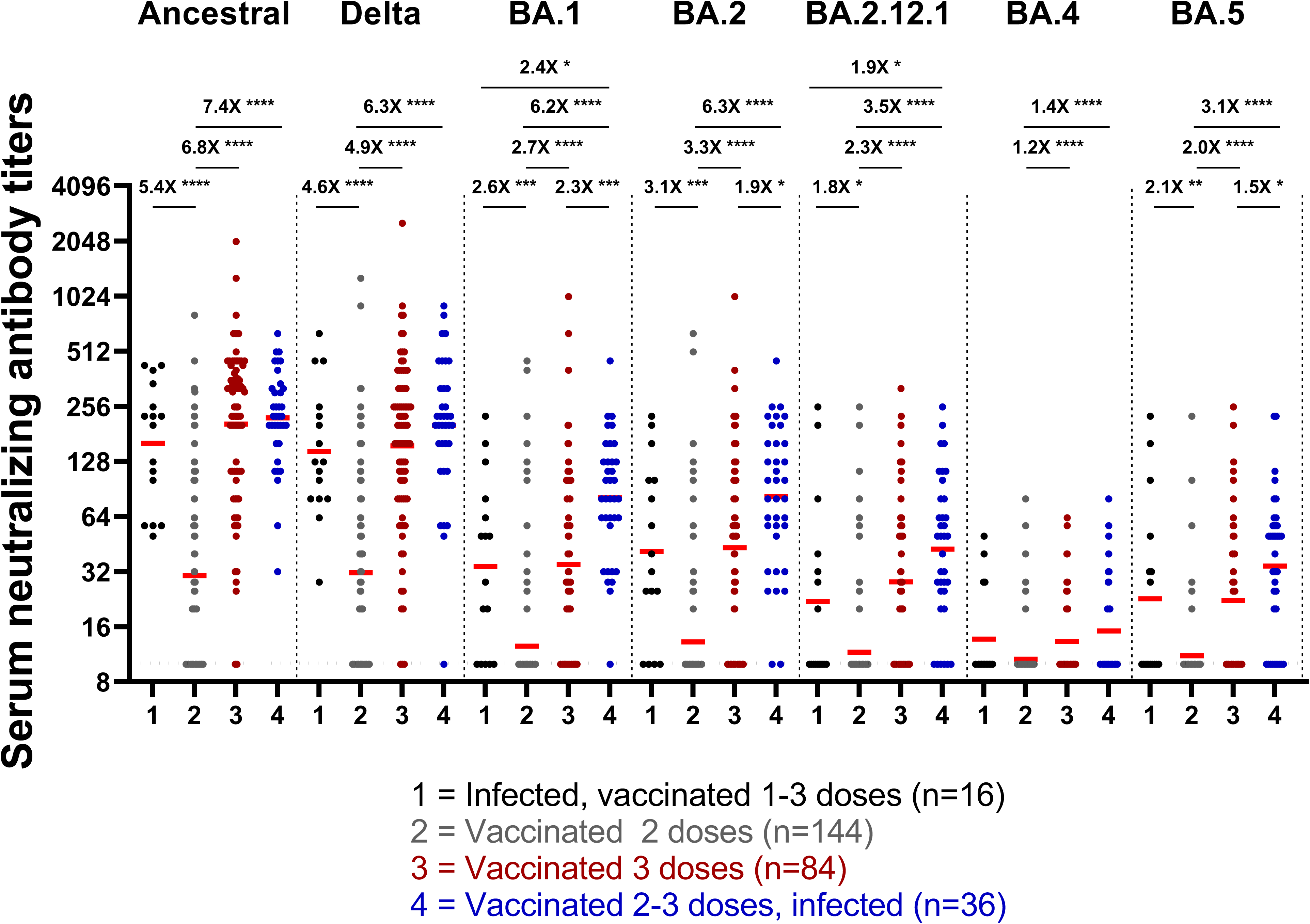
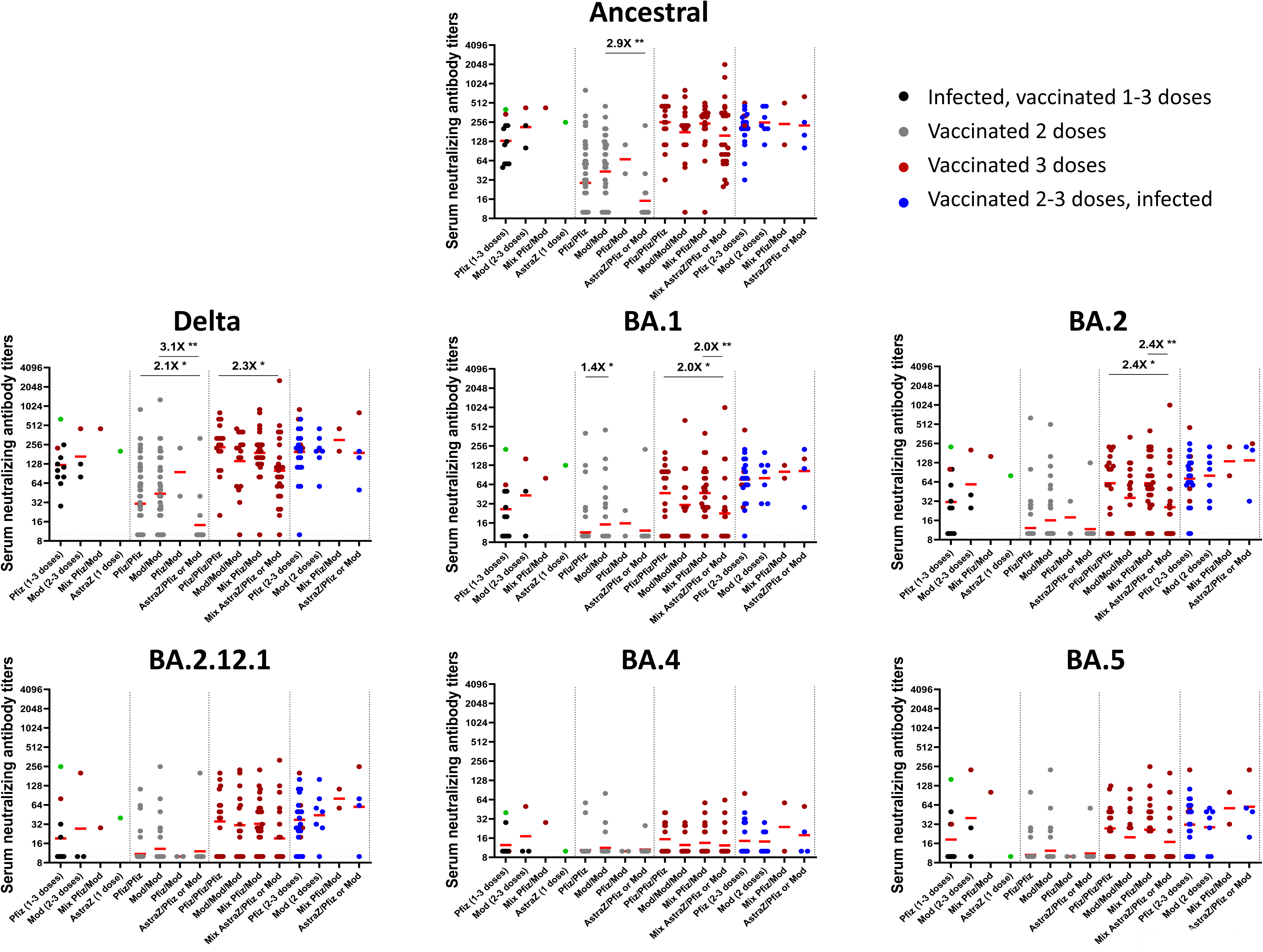
Neutralizing antibody titers against ancestral SARS-CoV-2, Delta, BA.1, BA.2, BA.2.12.1, BA.4 and BA.5 strains in all volunteers regarding the number of vaccine doses received and if they were infected before or after vaccinations. **(A)** Group 1 (n=16); infected, Pfizer (n=11), Moderna (n=3), Pfizer/Moderna (n=1) or AstraZeneca (n=1)); Groupe 2 (n=144); 2 doses of Pfizer (n=85), Pfizer/Moderna or vice versa (n=2), Moderna/Moderna (n=41) and AstraZeneca and Pfizer or Moderna (n=16); Group 3 (n=84); 3 doses of Pfizer (n=17), 3 doses of Moderna (n=16), Moderna/Moderna/Pfizer (n=5), Pfizer/Pfizer/Moderna (n=19), Pfizer/Moderna/Pfizer or Moderna (n=3), AstraZeneca/AstraZeneca/Pfizer or Moderna (n=4) or AstraZeneca/Pfizer or Moderna/Moderna (n=20); Group 4 (n=36): participants received at least 2 doses of vaccine (Pfizer/Pfizer (n=25), Moderna/Moderna (n=7), AstraZeneca/ AstraZeneca or AstraZeneca/Moderna (n=4)) and then were infected. **(B)** Neutralizing antibody titers for each participant against ancestral SARS-CoV-2, Delta, BA.1, BA.2, BA.2.12.1, BA.4 and BA.5 strains regarding type of vaccine regimen. Each circle represents a single participant. In the first group, the green and red points represent participants vaccinated with 1 and 3 doses and then infected, respectively. Bars identify geometric mean titers of the group. The horizontal dashed line indicates the limit of detection for the neutralization assay (neutralizing titer of 10). The samples that did not neutralize SARS-CoV-2 at 1:20 serum dilution was given a neutralizing titer of 10 for graphic representation and statistical analysis. The fold-change of the geometric mean titer is denoted on the line. Significance was assessed with Kruskal-Wallis one-way ANOVA followed by Dunn’s multiple comparison test (\**p* < 0.05; ** *p* < 0.01; *** *p* < 0.001; **** *p* < 0.0001).

## DISCUSSION

One of the important roles of vaccination is to generate broad, long-lasting immunity that will contribute to protect individuals from future infections or at least from severe clinical outcomes if infected. Since the emergence of the Omicron lineage in late 2021, several Omicron variants continue to evolve into new subvariants which are increasingly resistant to monovalent and bivalent vaccination [21–24].

During the first two years of the COVID-19 pandemic (i.e from March 2020 to February 2022), the provincial government of Québec in Canada has used a plethora of measures and restrictions on its territory in an attempt to reduce the infection rate. These measures included house confinement, curfew, suspension of activities deemed non-essential, including within the manufacturing and service industry, and mandatory online class for school and university (no face-to-face). These measures varied in time and length and were generally implemented based on infection rate within sub-regions of the province. During this period, grocery stores were considered essential institutions in support of food and general supplies, and remained open most of the time. Public health measures (e.g., mask wearing, social distancing, staying home when sick, etc.) were enforced and generally well-respected. Hardware stores and restaurants were instead intermittently opened and closed by health authorities over the same period, and were at higher risk of SARS-CoV-2 transmission due to the intrinsically social nature of these businesses and difficulty to enforce public health measures. In December 2020, vaccines were approved in Canada and priority was given to essential workers as a measure to protect themselves, their families, co-workers, and their community, while ensuring the constant availability of food/hardware supplies.

Many studies have been performed to analyze the antibody response of health-care workers, school children and staff and nursing homes in Canada and elsewhere [12–14, 21, 23–25], however, no serological studies in a cohort of food/retail workers have been conducted in Canada.

Antibody measurements are a crucial aspect of estimating the level of herd immunity in communities. Importantly, NtAbs are important for virus clearance and their titers have been demonstrated to be correlated with vaccine efficacy [11, 26, 27]. Indeed, several studies have shown that high levels of NtAbs are associated with protection from symptomatic SARS-CoV-2 infection after vaccination [11, 28, 29], however, all the studies have different approaches to estimate the relationship between NtAb titers and vaccine efficacy. A recent study performed by Khoury et al. [26] has analyzed four phase 3 clinical studies evaluating the performance of COVID-19 vaccines against the ancestral SARS-CoV-2 strain at the beginning of the vaccine campaign and before the emergence of the VOCs. Each study [11, 28–30] reported on a significant relationship between neutralizing titers and vaccine efficacy using different neutralization methodologies. Interestingly, Khoury et al. found that when centering the data on the GMT elicited by the vaccine, the four studies converged on a common correlating prediction between neutralization capacity and protection against infection [26], supporting the use of NtAb titers to predict the efficacy of new vaccines or vaccine efficacy against new VOCs (when fold drop in NtAb titer for the variant can be estimated).

In this study we compared the neutralizing antibody levels against the ancestral SARS-CoV-2, Delta, Omicron BA.1, BA.2, BA.2.12.1, BA.4 and BA.5 strains in 280 serum samples collected from vaccinated food and retail workers including those from grocery and hardware stores, restaurants and bars recruited in the Québec City area (Capitale-Nationale and Chaudière-Appalaches administrative regions) during the emergence of Omicron BA.1 variant (October 2021 to May 2022). By the time of the sample collection, all participants had received at minimum one dose of either mRNA vaccines (BNT162b2, Pfizer-BioNTech); mRNA-1273, Moderna) or viral vector vaccine (ChAdOx1-S, AstraZeneca).

When we analyzed each of the participating groups of workers (grocery stores, restaurants/bars or hardware stores), no statistical difference was observed in the GMTs of the groups against any of the SARS-CoV-2 strains tested. Overall, when considering all participants, we found no statistical difference in the NtAb titers against the ancestral SARS-CoV-2 strain and the Delta variant. In general, the NtAb response was reduced against the other variants tested, where the GMTs against BA.1, BA2, BA.2.12.1, BA.4 and BA.5 were reduced by 3.3-, 3-, 4-, 6.3-, and 4.7-fold compared to the ancestral SARS-CoV-2, respectively. This decreasing pattern of NtAb response is in line with studies reported in other populations such as healthcare workers, healthy vaccinated adults and adolescents and immunocompromised patients [31–37]. No statistically significant difference in the GMTs was observed between male/female participants, age groups (<60 years versus ≥60 years) and in males and females with chronic disease, smoking or vaping or among those volunteers with all three characteristics. Interestingly, our study found a significant difference in the GMTs against all SARS-CoV-2 strains tested except Delta in obese females (BMI ≥30; n=48; median delay of 95 days between last vaccination/infection and samples collection for BMI≥30 versus 96 days for BMI<30). Of note, according to the vaccine safety and efficacy information for Pfizer, Moderna, and Johnson & Johnson formulations showed similar efficacy in individuals with or without obesity [38]. However, a systematic review [39] of published studies on the safety and efficacy of COVID-19 vaccine in people who were overweight or obese reported that in nine out of twelve studies a reduced response with increased BMI was observed. Importantly, contradictory results may be due to different measure of obesity (e.g. central obesity or BMI), vaccination, comorbidities, the use of antibody titer kits or assays, age, sex, measuring antibodies at varying time points, etc. Therefore, more research needs to be done to assess the impact of obesity on immunogenicity of SARS-CoV-2 vaccines.

The seroprevalence of ancestral SARS-CoV-2 antibodies in the 280 participants during the period of October 2021 to May 2022 (emergence of the first Omicron sub-lineage (BA.1) was 80%. Cross-variant neutralization capacity was found in 99%, 55%, 58%, 46%, 21% and 39% of the participants against the Delta, BA.1, BA.2, BA.2.12.1, BA.4 and BA.5, respectively. Similar to other studies in different groups of individuals [21, 31–37, 40], our data show that uninfected food and retail workers from Québec who received 2 doses of vaccine had significantly lower NtAb titers against the ancestral SARS-CoV-2 and all variants than those who were infected and then vaccinated (≥1 dose), vaccinated (≥2 doses) and then infected, or those who received 3 doses of vaccine. As previously reported, hybrid immunity led to better humoral response against SARS-CoV-2 variants than vaccination alone, as participants who were infected after two or three doses of vaccine had higher NtAbs levels against BA.1, BA.2 and BA.5 variants, compared with other groups, suggesting a substantial degree of cross-reactive natural immunity. Therefore, workers with hybrid immunity acquired during Omicron BA.1 emergence might be better protected against reinfection with subsequent Omicron variants such as BA.2, BA.4/5 etc.

Our study has several limitations. An important limitation is that the interval between the last vaccination/infection and sample collection was variable; for example, for participants who received 2 or 3 doses of vaccine, the delay was 157 and 48 days (median), respectively, amplifying the potential difference in the antibody response. Importantly, our conclusions concerning the differences between the 4 groups apply even when comparing GMTs for similar intervals. We were also unable to confirm the specific SARS-CoV-2 strain that caused the infection among participants. The use of different vaccine platforms finally highlights a potential limitation of our study as the immunogenicity of each vaccine type differs. Reassuringly, however, almost all participants (279/280) received at least one mRNA vaccine.

In summary, we assessed for the first time the neutralizing antibody response of a unique population of food and retail workers. Overall, we found that vaccination was associated with higher neutralizing activity against pre-Omicron variants and that vaccination followed by infection was associated with higher neutralizing activity against Omicron sub-lineages, in line with the humoral response in other populations (health-care workers, healthy vaccinated adults and adolescents as well as immunocompromised patients). Interestingly, we did not observe any significant difference in the NtAb response in terms of sex, age, or male/female with chronic diseases, smokers or vaping or among those volunteers with all three characteristics. Additional public health measures may be warranted to increase antibody response against new SARS-CoV-2 variants such as updated vaccines for the population.

## Data Availability

All data produced in the present work are contained in the manuscript

## Supplementary Data

Supplementary material associated with this article can be found in the online.

## Declaration of competing interest

The authors declared no competing interests.

## Funding

This study was funded by the Public Health of Canada through the COVID-19 Immunity Task Force (CITF) to CG, JP, JFM, DB, ST and MB. This research was supported in part by the Sentinel North Research Chair at Université Laval (funded by the Canada First Research Excellence Fund) and by the Canada Research Chair to M.B.

## Ethical approval

This study was approved by the « Comité d’éthique de la recherche du CHU de Québec-Université Laval (registration number 2021-5744). A unique, anonymized identifier was assigned to each participant and used to store the data and the samples.

## Acknowledgements

The authors sincerely acknowledge the nurses and research coordinators at Centre de recherche du centre hospitalier universitaire de Québec for their involvement in this study and all donors who agreed to participate in this research project. We are grateful to the National Microbiology Laboratory (NML), Public Health Canada, for providing SARS-CoV-2 isolates (SARS-CoV-2 VOC B.1.617.2 (Delta), B1.1.529 (Omicron sub-lineage BA.1) and Omicron sub-lineages BA.2, BA.2.12.1, BA.4 and BA.5 used in this study.

## Author contributions

H.R. and M.B. designed the study. K.S. and S.T. wrote the clinical protocol and recruited the participants. H.R., M.S., M.G. and M.B. performed the experiments. M.B. supervised the study. H.R. and M.B. analyzed the data and wrote the manuscript. C.G., K.S., M.T., N.B., J.F.M., J.P., D.B., and S.T. revised the manuscript and provided scientific input. All authors have read and approved the final manuscript.

---

**Supplementary Figure 1**

Neutralizing antibody titers after the most recent vaccine dose or infection and the sample collection. Time was grouped into 7-30 days, 31-60 days, 61-90 days and over 90 days. **(A)** Groupe 1, infected, then vaccinated with ≥ 1 dose. **(B)** Groupe 2, vaccinated with 2 doses **(C)** Groupe 3, vaccinated with 3 doses. **(D)** Groupe 4, vaccinated with ≥ 2 dose and infected. Significance was assessed with Kruskal-Wallis one-way ANOVA followed by Dunn’s multiple comparison test (\*\**p* < 0.01; \*\*\**p* < 0.001).

**Supplementary Figure 2**

Neutralizing antibody titers against ancestral SARS-CoV-2, Delta, BA.1, BA.2, BA.2.12.1, BA.4 and BA.5 strains for each of the groups: (grocery stores (n=105), hardware stores (n=42) and restaurant/bars (n=133). Each circle represents a single participant. Bars identify geometric mean titers of the group. The horizontal dashed line indicates the limit of detection for the neutralization assay (neutralizing titer of 10). The samples that did not neutralize SARS-CoV-2 at 1:20 serum dilution was given a neutralizing titer of 10 for graphic representation and statistical analysis. The fold-change of the geometric mean titer is denoted on the line. Significance was assessed with Kruskal-Wallis one-way ANOVA followed by Dunn’s multiple comparison test (\**p* < 0.05; \*\**p* < 0.01; \*\*\**p* < 0.001; \*\*\*\**p* < 0.0001).

**Supplementary Figure 3**

Neutralizing antibody titers against ancestral SARS-CoV-2, Delta, BA.1, BA.2, BA.2.12.1, BA.4 and BA.5 strains in male and female with BMI ≥30, chronic disease, smokers or vaping or among those volunteers with all three characteristics. Each circle represents a single participant. Bars identify geometric mean titers of the group. The horizontal dashed line indicates the limit of detection for the neutralization assay (neutralizing titer of 10). The samples that did not neutralize SARS-CoV-2 at 1:20 serum dilution were given a neutralizing titer of 10 for graphic representation and statistical analysis. Significance was assessed with Kruskal-Wallis one-way ANOVA followed by Dunn’s multiple comparison test (\**p*< 0.05; \*\**p* < 0.01).

## Notes

### Competing Interest Statement

The authors have declared no competing interest.

### Funding Statement

This study was funded by Public Health of Canada through the COVID‐19 Immunity Task Force (CITF) to CG, JP, JFM, DB, ST and MB and the Sentinel North Research Chair at Universite Laval (funded by the Canada First Research Excellence Fund) as well as the Canada Research Chair to M.B.

### Author Declarations

CHU de Quebec-Universite Laval Research Ethics Board (registration number 2021-5744)

## References

1. Singh J, Rahman SA, Ehtesham NZ, Hira S, Hasnain SE. SARS-CoV-2 variants of concern are emerging in India. Nat Med 2021; 27(7): 1131–3.

2. Liu C, Ginn HM, Dejnirattisai W, et al. Reduced neutralization of SARS-CoV-2 B.1.617 by vaccine and convalescent serum. Cell 2021; 184(16): 4220–36 e13.

3. Tian D, Sun Y, Zhou J, Ye Q. The Global Epidemic of the SARS-CoV-2 Delta Variant, Key Spike Mutations and Immune Escape. Front Immunol 2021; 12: 751778.

4. Torjesen I. Covid-19: Omicron may be more transmissible than other variants and partly resistant to existing vaccines, scientists fear. BMJ 2021; 375: n2943.

5. VanBlargan LA, Errico JM, Halfmann PJ, et al. An infectious SARS-CoV-2 B.1.1.529 Omicron virus escapes neutralization by therapeutic monoclonal antibodies. Nat Med 2022.

6. World Health Organization (WHO). Tracking SARS-CoV-2 variants. https://www.who.int/activities/tracking-SARS-CoV-2-variants. .

7. Kurhade C, Zou J, Xia H, et al. Low neutralization of SARS-CoV-2 Omicron BA.2.75.2, BQ.1.1, and XBB.1 by parental mRNA vaccine or a BA.5-bivalent booster. Nat Med 2022.

8. Addetia A, Crawford KHD, Dingens A, et al. Neutralizing Antibodies Correlate with Protection from SARS-CoV-2 in Humans during a Fishery Vessel Outbreak with a High Attack Rate. J Clin Microbiol 2020; 58(11).

9. McMahan K, Yu J, Mercado NB, et al. Correlates of protection against SARS-CoV-2 in rhesus macaques. Nature 2021; 590(7847): 630-4.

10. Hobson D, Curry RL, Beare AS, Ward-Gardner A. The role of serum haemagglutination-inhibiting antibody in protection against challenge infection with influenza A2 and B viruses. J Hyg (Lond) 1972; 70(4): 767–77.

11. Khoury DS, Cromer D, Reynaldi A, et al. Neutralizing antibody levels are highly predictive of immune protection from symptomatic SARS-CoV-2 infection. Nat Med 2021; 27(7): 1205–11.

12. Science M, Bolotin S, Silverman M, et al. SARS-CoV-2 antibodies in Ontario health care workers during and after the first wave of the pandemic: a cohort study. CMAJ Open 2021; 9(4): E929–E39.

13. Eldesoukey N, Gaafar T, Enein AA, et al. SARS-CoV-2 antibody seroprevalence rates among Egyptian blood donors around the third wave: Cross-sectional study. Health Sci Rep 2022; 5(3): e634.

14. Lewin A, De Serres G, Gregoire Y, et al. Seroprevalence of SARS-CoV-2 antibodies among blood donors in Quebec: an update from a serial cross-sectional study. Can J Public Health 2022; 113(3): 385–93.

15. Zinszer K, McKinnon B, Bourque N, et al. Seroprevalence of SARS-CoV-2 Antibodies Among Children in School and Day Care in Montreal, Canada. JAMA Netw Open 2021; 4(11): e2135975.

16. West EA, Kotoun OJ, Schori LJ, et al. Seroprevalence of SARS-CoV-2 antibodies, associated factors, experiences and attitudes of nursing home and home healthcare employees in Switzerland. BMC Infect Dis 2022; 22(1): 259.

17. Kim Santerre, Mathieu Thériault, Nicholas Brousseau, Samuel Rochette, Joelle Pelletier, Caroline Gilbert, Jean-Francois Masson, Mariana Baz, Denis Boudreau, Sylvie Trottier. Cohort Profile: Prospective Cohort to Study the COVID-19 Immune Response in Retail Workers in Quebec, Canada (CISACOV). medRxiv 2023.08.18.23294172; doi: 10.1101/2023.08.18.23294172.

18. Baz M, Boonnak K, Paskel M, et al. Nonreplicating influenza A virus vaccines confer broad protection against lethal challenge. MBio 2015; 6(5): e01487–15.

19. Baz M, Paskel M, Matsuoka Y, et al. Replication and immunogenicity of swine, equine, and avian h3 subtype influenza viruses in mice and ferrets. J Virol 2013; 87(12): 6901–10.

20. Reed LJ, and H. Muench. . A simple method of estimating fifty percent endpoints. Am. J. Hyg. 27:493–497. 1938.

21. Qu P, Faraone J, Evans JP, et al. Neutralization of the SARS-CoV-2 Omicron BA.4/5 and BA.2.12.1 Subvariants. N Engl J Med 2022; 386(26): 2526–8.

22. Wang Q, Guo Y, Iketani S, et al. Antibody evasion by SARS-CoV-2 Omicron subvariants BA.2.12.1, BA.4 and BA.5. Nature 2022; 608(7923): 603–8.

23. Kurhade C, Zou J, Xia H, et al. Low neutralization of SARS-CoV-2 Omicron BA.2.75.2, BQ.1.1 and XBB.1 by parental mRNA vaccine or a BA.5 bivalent booster. Nat Med 2022.

24. Wang Q, Bowen A, Valdez R, et al. Antibody Response to Omicron BA.4-BA.5 Bivalent Booster. N Engl J Med 2023.

25. Ionescu IG, Skowronski DM, Sauvageau C, et al. BNT162b2 effectiveness against Delta and Omicron variants of SARS-CoV-2 in adolescents aged 12-17 years, by dosing interval and duration. J Infect Dis 2023.

26. Khoury DS, Schlub TE, Cromer D, et al. Correlates of Protection, Thresholds of Protection, and Immunobridging among Persons with SARS-CoV-2 Infection. Emerg Infect Dis 2023; 29(2): 381–8.

27. Carazo S, Skowronski DM, Brisson M, et al. Protection against omicron (B.1.1.529) BA.2 reinfection conferred by primary omicron BA.1 or pre-omicron SARS-CoV-2 infection among health-care workers with and without mRNA vaccination: a test-negative case-control study. Lancet Infect Dis 2023; 23(1): 45–55.

28. Feng S, Phillips DJ, White T, et al. Correlates of protection against symptomatic and asymptomatic SARS-CoV-2 infection. Nat Med 2021; 27(11): 2032–40.

29. Gilbert PB, Montefiori DC, McDermott AB, et al. Immune correlates analysis of the mRNA-1273 COVID-19 vaccine efficacy clinical trial. Science 2022; 375(6576): 43–50.

30. Bergwerk M, Gonen T, Lustig Y, et al. Covid-19 Breakthrough Infections in Vaccinated Health Care Workers. N Engl J Med 2021; 385(16): 1474–84.

31. Wang B, Xu J, Wu S, et al. Comparative characterization of antibody responses induced by Ad5-vectored spike proteins of emerging SARS-CoV-2 VOCs. Signal Transduct Target Ther 2022; 7(1): 257.

32. Banerjee A, Lew J, Kroeker A, et al. Immunogenicity of convalescent and vaccinated sera against clinical isolates of ancestral SARS-CoV-2, Beta, Delta, and Omicron variants. Med (N Y) 2022; 3(6): 422–32 e3.

33. Mu X, Cohen CA, Leung D, et al. Antibody and T cell responses against wild-type and Omicron SARS-CoV-2 after third-dose BNT162b2 in adolescents. Signal Transduct Target Ther 2022; 7(1): 397.

34. Cheng SMS, Mok CKP, Leung YWY, et al. Neutralizing antibodies against the SARS-CoV-2 Omicron variant BA.1 following homologous and heterologous CoronaVac or BNT162b2 vaccination. Nat Med 2022; 28(3): 486–9.

35. Rosa Duque JS, Wang X, Leung D, et al. Immunogenicity and reactogenicity of SARS-CoV-2 vaccines BNT162b2 and CoronaVac in healthy adolescents. Nat Commun 2022; 13(1): 3700.

36. Ferreira VH, Solera JT, Hu Q, et al. Homotypic and heterotypic immune responses to Omicron variant in immunocompromised patients in diverse clinical settings. Nat Commun 2022; 13(1): 4489.

37. Keppler-Hafkemeyer A, Greil C, Wratil PR, et al. Potent high-avidity neutralizing antibodies and T cell responses after COVID-19 vaccination in individuals with B cell lymphoma and multiple myeloma. Nat Cancer 2023; 4(1): 81–95.

38. Nasr MC, Geerling E, Pinto AK. Impact of Obesity on Vaccination to SARS-CoV-2. Front Endocrinol (Lausanne) 2022; 13: 898810.

39. Fu C, Lin N, Zhu J, Ye Q. Association between Overweight/Obesity and the Safety and Efficacy of COVID-19 Vaccination: A Systematic Review. Vaccines (Basel) 2023; 11(5).

40. Hachmann NP, Miller J, Collier AY, et al. Neutralization Escape by SARS-CoV-2 Omicron Subvariants BA.2.12.1, BA.4, and BA.5. N Engl J Med 2022; 387(1): 86–8.

