## Supplementary figures and images for "Live virus neutralizing antibodies against pre and post Omicron strains in food and retail workers in Québec, Canada"

### Supplementary figures 1-3

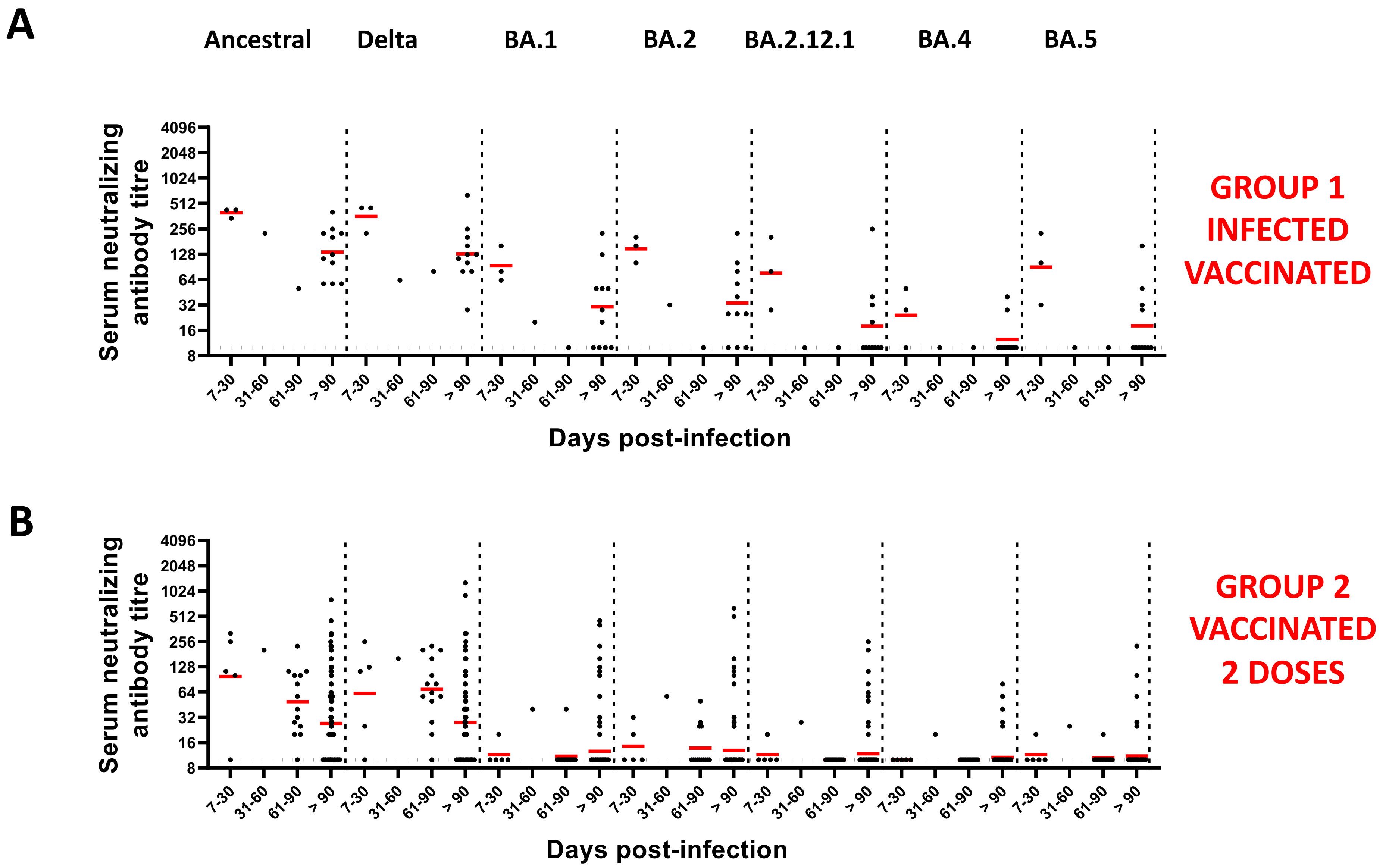


Supplementary Figure 1

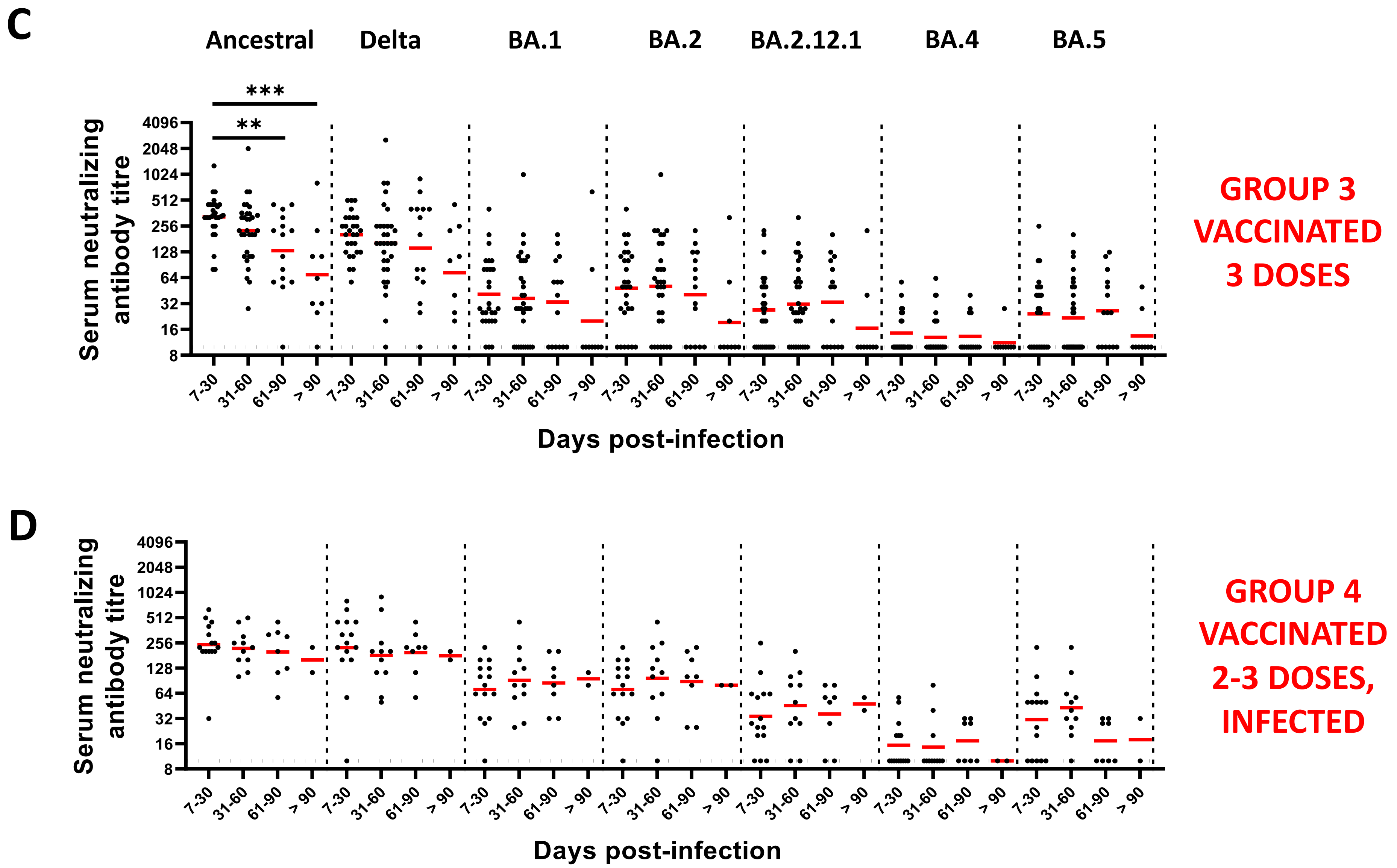


Supplementary Figure 1

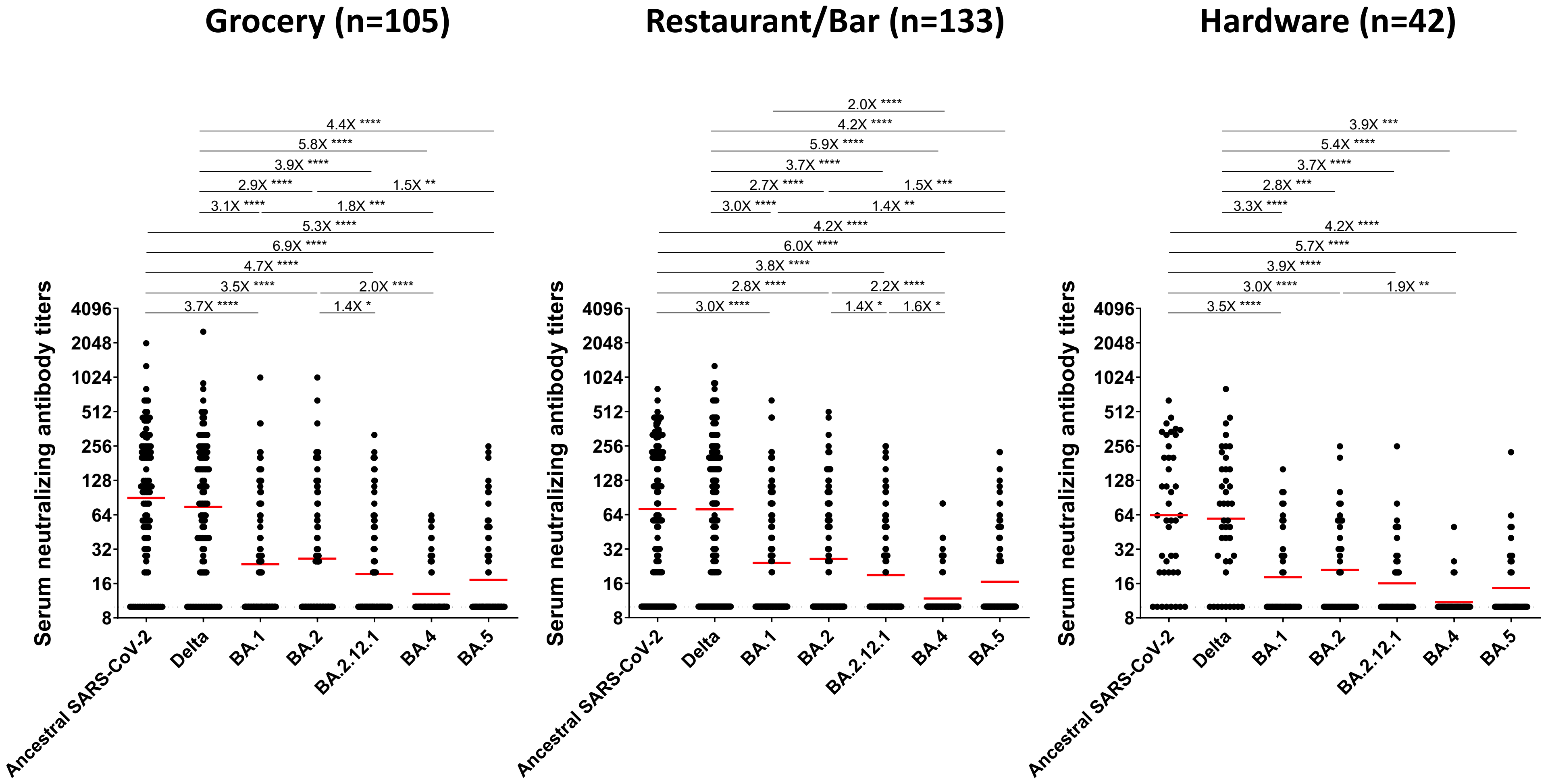


**Supplementary Figure 2**

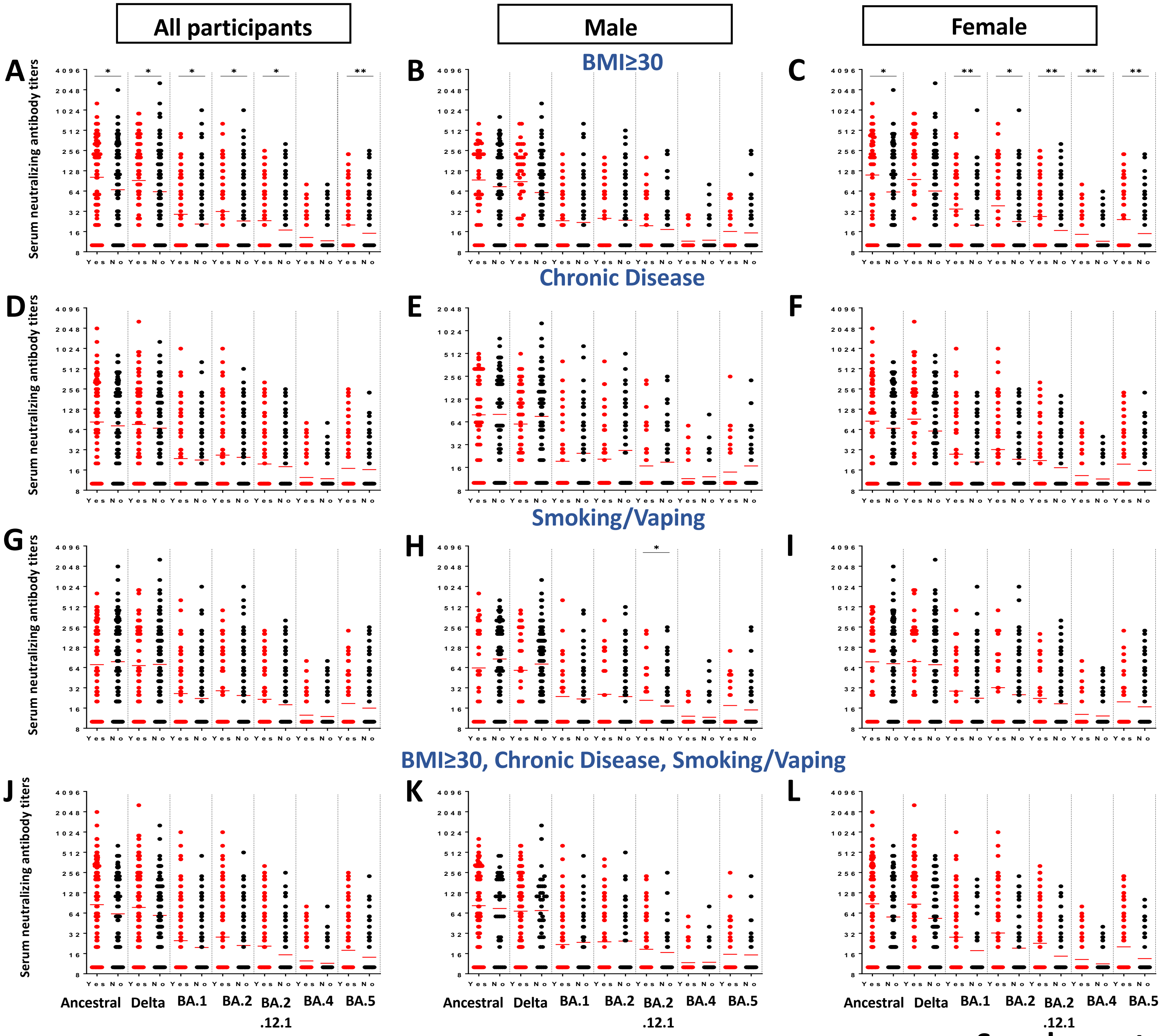


Supplementary Figure 3
